# Childhood trauma in patients with epileptic versus non-epileptic seizures

**DOI:** 10.1101/2022.08.16.22278830

**Authors:** Tianren Yang, Caitlin Roberts, Toby Winton-Brown, Michael Lloyd, Patrick Kwan, Terence J O’Brien, Dennis Velakoulis, Genevieve Rayner, Charles B Malpas

## Abstract

**Objective:** Childhood trauma has been implicated as a risk factor for the aetiology of psychogenic non-epileptic seizures (PNES). Relatively little attention has been paid to whether profiles of specific trauma types differ between patients with epilepsy and PNES. Investigating childhood trauma profiles in these patient groups may identify psychological vulnerabilities the predispose to developing PNES, and aid early diagnoses, prevention, and treatment.

**Methods:** Data were collected from two cohorts (*n*_*Retrospective*_=203; *n*_Prospective_=209) admitted to video-EEG monitoring units in Melbourne Australia. The Childhood Trauma Questionnaire domain score differences between patient groups were investigated using standardised effect sizes and general linear mixed-effects models (GLMMs). Receiver operating characteristic curves were used to investigate classification accuracy.

**Results:** In the retrospective cohort, patients diagnosed with PNES reported greater childhood emotional abuse, emotional neglect, physical abuse, sexual abuse, and physical neglect relative to epilepsy patients. These differences were replicated in the prospective cohort, except for physical abuse. GLMMs revealed significant main effects for group in both cohorts, but no evidence for any group by domain interactions. Reported sexual abuse showed the best screening performance of PNES, although no psychometric scores were adequate as isolated measures.

**Significance:** Patients with PNES report greater frequency of childhood trauma than patients with epilepsy. This effect appears to hold across all trauma types, with no strong evidence emerging for a particular trauma type that is more prevalent in PNES. From a practical perspective, inquiring regarding a history of sexual abuse shows the most promise as a screening measure.

**Summary:**

- Relatively little attention has been paid to whether profiles of specific trauma types differ between patients with epilepsy and PNES.
- We collected self-reported childhood trauma information from two independent cohorts of patients undergoing VEM
- Patients with PNES report greater frequency of childhood trauma than patients with epilepsy.
- This effect appears to hold across all trauma types, with no strong evidence emerging for a particular trauma type that is more prevalent in PNES.
- Inquiring regarding a history of sexual abuse shows the most promise as a screening measure.

## Introduction

Psychogenic nonepileptic seizures (PNES) are commonly misdiagnosed as epileptic seizures, but on video-EEG monitoring (VEM) lack accompanying epileptiform EEG activity, have semiological features distinct from epileptic seizures, and are underpinned by psychological factors (1). Due to the similar presentations, diagnosing PNES is challenging, with an average diagnostic delay of seven to 16 years after seizure onset (2, 3). Investigating PNES risk factors may facilitate early accurate diagnoses and decrease misguided use of limited health care resources (4) and other health burdens (5).

Modern theories emphasise that the aetiology of PNES is multi-factorial and involves complex interactions between environmental, genetic, and psychological factors, with experiences of trauma being the most common risk factor (6-9). Approximately 90% of people with PNES report having experienced traumatic events across their lifetime compared to 74.9% in the general population and 85% in epilepsy patients (1, 10). Specifically, childhood trauma increases an individual’s odds of developing PNES more than the same type of trauma occurring in adulthood (11).

Analysing childhood trauma experience in PNES may inform PNES aetiology and treatment planning. Early studies found higher rates of childhood sexual and/or physical abuse in people with PNES compared to those with epilepsy (12), although subsequent findings have been more equivocal (1, 10, 13, 14). Recent studies assessing childhood trauma more comprehensively report increased odds of PNES diagnosis compared to epilepsy in people who experienced physical abuse and neglect (2.20 and 2.84, respectively) (13) and childhood psychological abuse (2.39) (14), but not other trauma types. Similarly, two studies found that patients with PNES experience significantly more severe overall childhood trauma, with emotional abuse and neglect as the most potent trauma vulnerability factors for developing PNES (15, 16). In contrast, no significant differences were found in trauma experience between these patient groups when using gold standard VEM diagnostic criteria (1), although this study may have lacked statistical power to detect profile differences (n_PNES_ = 12, n_epilpesy_ = 61). PNES is a heterogeneous condition and there is growing evidence that psychological trauma might represent an aetiological factor in one or more subtypes of patients (17, 18).

These discrepancies of childhood trauma differences between PNES and epilepsy in the current literature may largely be explained by methodological limitations such as using smaller samples and non-gold standard diagnostic approaches to classify patients. Our study is the first to investigate childhood trauma profiles in large samples of patients with confirmed PNES and epilepsy and attempt to replicate the results in a separate cohort. We aim to compare childhood trauma profiles in patients diagnosed on in-patient video-EEG monitoring with PNES versus epilepsy, and to additionally examine the clinical utility of childhood trauma to identify those at risk of PNES.

## Methods

### Retrospective discovery cohort

A retrospective discovery cohort comprised patients who were admitted to VEM unit at The Royal Melbourne Hospital in Melbourne, Australia, between 2014 and 2017. Following an audit of clinical records, patients were included if they were aged 18 years or over and had completed the Child Trauma Questionnaire (CTQ), which was administered as part of routine clinical practice. Patients were not asked to complete the CTQ if they had an intellectual disability or other cognitive impairments severe enough to impair their understanding of the questionnaires.

### Prospective validation cohort

A prospective validation cohort included patients who were admitted to VEM units at The Royal Melbourne and The Alfred Hospitals in Melbourne, Australia, between April 2018 and October 2019. Inclusion and exclusion criteria were the same as for the retrospective cohort. Detailed clinical and psychometric data were collected as described below.

### Standard Protocol Approvals, Registrations, and Patient Consents

This study received approval from The Melbourne Health, The Alfred Health Research and Ethics Committee, and The University of Melbourne Human Ethics Advisory Group.

### Diagnoses

All patients were assessed by a multidisciplinary team consisting of neurologists, neuroradiologists, neuropsychiatrists, and neuropsychologists to reach a consensus diagnosis at the conclusion of the VEM admission (19). The patient’s clinical history, neurological examination, video-EEG recordings, neuroimaging (where applicable), neuropsychiatric, and neuropsychological assessments were considered in the diagnostic formulation, and patients were classified as one of the following: epileptic seizures (ES); psychogenic nonepileptic seizures (PNES); concomitant PNES and ES (PNES+ES); other non-epileptic events (ONE) such as cardiovascular events, vasovagal syncope, panic attacks; or non-diagnostic (NDX). For patients with ES and PNES+ES, the VEM results provided further classifications of focal or generalised epilepsy, temporal lobe or extra-temporal focal epilepsy, and seizure lateralisation (left, right, or bilateral). Patients with both PNES+ES were excluded from analyses in order to compare two robust diagnostic groups.

### Clinical Characteristics

Patient age, gender, current antiseizure medications (ASMs), disease duration, and event frequency were recorded at the time of admission. Typical event frequency was rated on a 13-point scale (from 0 – ‘seizure-free, off ASMs’ to 12 – ‘status epilepticus’) as described previously (20).

### Psychiatric Diagnoses

All patients underwent a specialist neuropsychiatric clinical assessment during the VEM admissions. Psychiatric diagnoses of anxiety and depressive disorders were based on the Diagnostic and Statistical Manual – Fifth Edition (21), and included current generalised anxiety disorder, post-traumatic stress disorder, panic disorder, and specific phobia. Patients were characterised with comorbid depressive disorders if they had any current major depressive disorder, dysthymia, or adjustment disorder.

### Psychological questionnaires

All questionnaires were administered as part of the standard comprehensive patient care. Patients in both cohorts completed the CTQ and the Neuropsychiatry Unit Cognitive Assessment (NUCOG). Patients in the prospective cohort also completed the Generalised Anxiety Disorders 7-Item (GAD-7), the Neurological Disorders Depression Inventory for Epilepsy (NDDI-E), the Wessex Dissociation Scale (WDS), and the Personality Inventory for DSM-5—Adult (PID-5). These questionnaires were chosen for their ease of administration and reliability in clinical populations. Full details of these instruments are included below.

### Childhood Trauma Questionnaire (CTQ)

The CTQ is a validated 28-item questionnaire assessing the severity of trauma in five domains: emotional abuse, emotional neglect, physical abuse, physical neglect, and sexual abuse (22). Patients are asked to consider their childhood experiences and rate the severity of traumatic experience described in each item on a five-point, Likert-type scale. The total score (out of 125) and five domain scores (each out of 25) correlate with trauma severity, with higher scores indicating more significant trauma. The CTQ also incorporates a minimisation scale that indicates the tendency to underreport childhood maltreatment.

### Neuropsychiatry Unit Cognitive Assessment (NUCOG)

The NuCOG was developed as a cognitive screening tool that consists of five cognitive domains: attention, visuoconstruction, memory, executive functions, and language (23). The NuCOG total score ranges between 0 to 100, with higher scores indicating less impaired cognitive function and was found to be reliable and valid in reflecting the cognitive function of the participants. NuCOG total score has been found to have high reliability and low measurement error in patients undergoing VEM (24, 25).

### Generalised Anxiety Disorders 7-Item (GAD-7)

The GAD-7 is a seven-item inventory with a four-point, Likert scale (rated from “0 – not at all” to “3 – nearly every day”) that assesses the severity of anxiety symptomatology over the past two weeks (26). The total summed score ranges from 0 to 21 with higher scores indicating greater symptomatology. The GAD-7 has been shown to be reliable and valid in measuring anxiety levels in patients with epilepsy (27) and has been widely used among patients with PNES (28).

### Neurological Disorders Depression Inventory for Epilepsy (NDDI-E)

The NDDI-E is a six-item inventory that was specifically developed to screen for major depressive disorder (MDD) in patients with epilepsy (29). Participants self-report their experience of depressive symptoms in the past two weeks on a four-point, Likert scale (rated from “1 – never” to “4 – always or often”). The total sum score ranges from 6 to 24 with higher scores indicating greater symptomatology.

### Wessex Dissociation Scale (WDS)

The WDS is a 40-item questionnaire with a six-point, Likert scale (rated from “1 – never” to “6 – all the time”) that assesses psychopathology related dissociative symptoms (30). Higher scores indicate greater levels of dissociation, with total score ranging between 40 and 240.

### Personality Inventory for DSM-5—Adult (PID-5)

The PID-5 is a 220-item inventory developed to provide a dimensional measure of maladaptive personality traits (31). It is designed to to assess the emerging model for personality disorders based on personality dysfunction and pathological personality traits outlined in DSM-5. The PID-5 consists of the following five domains: negative affect, detachment, antagonism, disinhibition, and psychoticism. Participants self-report how much each item describes themselves on a four-point, Likert scale (rated from “0 – very false or often false” to “4 – very true or often true”). The psychometric properties of the PID-5 have been well established across a range of conditions. For example, the Cronbach’s (32) α is high for the negative affect (α = .93), detachment (α = .96), antagonism (α = .94), disinhibition (α = .84), and psychoticism (α = .96) domain scores (33). Relatively little evidence has investigated the temporal stability of the PID-5, however there is some evidence that the change over 1-2 years is relatively small across scores (median *d* = -0.12) and the rank order stability estimate is acceptable (median *r* = .68).(34). In terms of validity, the PID-5 shows good correspondence to semi-structured clinical interviews, which supports the use of the PID-5 as an adjunct to clinical diagnosis (35). Relatively few studies have investigated the PID-5 in epilepsy populations, however there is emerging evidence of different PID-5 profiles in PNES patients compared to those with epilepsy (36). PID-5 scores are also predictive of mis-diagnosis when using screening measures for depression and anxiety in patients undergoing VEM (37).

### Data Availability

Anonymised data, and the standardized proforma used to extract information on patient demographics and clinical characteristics, will be shared by request from any qualified investigator.

### Statistical analyses

Mean differences between the PNES and epilepsy groups on the CTQ were first investigated by computing standardised effect sizes, in the form of Hedges’ *g*, first for the retrospective cohort then for the prospective cohort. Effect sizes were interpreted using Cohen’s rules of thumb (38) for small (*g* <= 0.50), medium (>0.50 *g* <= 0.80), and large (*g* > 0.80). An effect size was considered successfully replicated when: (1) the point estimate computed in the prospective cohort was captured by the confidence intervals computed in the retrospective cohort, and (2) the confidence intervals did not capture zero in both the retrospective and prospective cohort. Profile differences between patients with epilepsy and PNES were then investigated using general linear mixed-effects models (GLMMs). CTQ domain score was entered as the dependent variable with age at admission, gender, CTQ domain, and the VEM diagnostic group (epilepsy versus PNES) entered as predictors. A random intercept was specified for each participant. All continuous variables (include CTQ domain scores) were centred and scaled prior to analysis. Robust standard errors and confidence intervals were computed using bootstrapping with 2000 replicates. Effect sizes, in the form of ω^2^_p_, were used to determine statistically supported effects.

The associations between psychometric markers of psychopathology, continuous clinicodemographic variables in the prospective cohort, and CTQ scores were examined using Spearman’s correlation. Robust confidence intervals were computed via bootstrapping. Generalised linear models (GZLMs) were computed to investigate the association between CTQ scores and diagnostic group (PNES versus epilepsy) over and above clinicodemographic variables. Each CTQ domain score was entered into the baseline model and the Akaike Information Criteria (AIC) and likelihood ratio test were inspected to determine if the model fit was substantially improved. Finally, receiver operating characteristic curves (ROC) were computed to investigate the classification performance of the raw CTQ scores. The optimal cut-off was determined using Youden’s *J* (39) statistic in the retrospective cohort and then applied to the prospective cohort to compute classification performance metrics. All analyses were performed using the R software (40).

## Results

### Sample characteristics

Of the 203 patients in the retrospective discovery cohort, the majority were diagnosed with epilepsy (*n* = 144, 71%) compared to PNES (*n* = 59, 29%). Of the patients with epilepsy, the majority had focal epilepsy (*n* = 120, 84%), followed by generalised epilepsy (*n* = 21, 15%), and mixed focal and generalised (*n* = 2, 1%). There were more females (*n* = 126, 62%) compared to males (*n* = 77, 38%). Of the 209 patients in the prospective validation cohort, the majority were diagnosed with epilepsy (*n* = 172, 82%) compared to PNES (*n* = 37, 18%). The most common epilepsy diagnosis was focal epilepsy (*n* = 145, 86%), followed by generalised (*n* = 23, 14%), and combined focal and generalised (*n* = 1). There were more females (*n* = 116, 56%) compared to males (*n* = 93, 44%). Overall, there were more females in the PNES groups relatively to the epilepsy groups. In the retrospective cohort, 51% of epilepsy patients were female compared to 88% of PNES patients (χ^2^(1) = 24.00, *p* < .001). A less pronounced imbalance was observed in the prospective cohort, with 52% of epilepsy patients being female compared to 70% of PNES patients (χ^2^ (1) = 3.97, *p* = .05). Further clinicodemographic information is shown in Table 1.

**Table 1.**
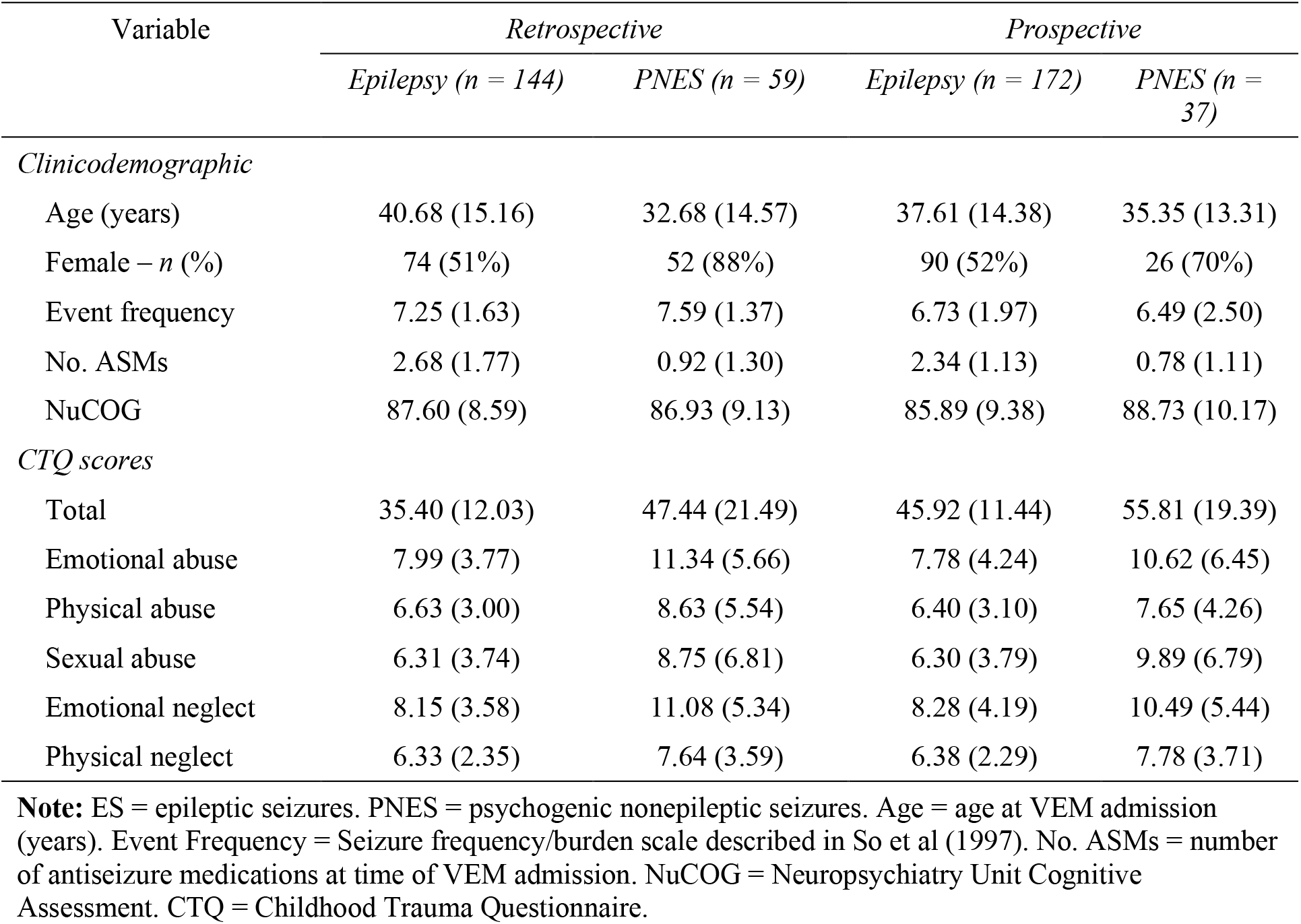
Sample characteristics for the retrospective and prospective cohorts – M (SD)

| Variable | *Retrospective* |  | *Prospective* |  |
| --- | --- | --- | --- | --- |
|  | *Epilepsy (n = 144)* | *PNES (n = 59)* | *Epilepsy (n = 172)* | *PNES (n = 37)* |
| *Clinicodemographic* |  |  |  |  |
| Age (years) | 40.68 (15.16) | 32.68 (14.57) | 37.61 (14.38) | 35.35 (13.31) |
| Female – n (%) | 74 (51%) | 52 (88%) | 90 (52%) | 26 (70%) |
| Event frequency | 7.25 (1.63) | 7.59 (1.37) | 6.73 (1.97) | 6.49 (2.50) |
| No. ASMs | 2.68 (1.77) | 0.92 (1.30) | 2.34 (1.13) | 0.78 (1.11) |
| NuCOG | 87.60 (8.59) | 86.93 (9.13) | 85.89 (9.38) | 88.73 (10.17) |
| *CTQ scores* |  |  |  |  |
| Total | 35.40 (12.03) | 47.44 (21.49) | 45.92 (11.44) | 55.81 (19.39) |
| Emotional abuse | 7.99 (3.77) | 11.34 (5.66) | 7.78 (4.24) | 10.62 (6.45) |
| Physical abuse | 6.63 (3.00) | 8.63 (5.54) | 6.40 (3.10) | 7.65 (4.26) |
| Sexual abuse | 6.31 (3.74) | 8.75 (6.81) | 6.30 (3.79) | 9.89 (6.79) |
| Emotional neglect | 8.15 (3.58) | 11.08 (5.34) | 8.28 (4.19) | 10.49 (5.44) |
| Physical neglect | 6.33 (2.35) | 7.64 (3.59) | 6.38 (2.29) | 7.78 (3.71) |

**Note:** ES = epileptic seizures. PNES = psychogenic nonepileptic seizures. Age = age at VEM admission (years). Event Frequency = Seizure frequency/burden scale described in So et al (1997). No. ASMs = number of antiseizure medications at time of VEM admission. NuCOG = Neuropsychiatry Unit Cognitive Assessment. CTQ = Childhood Trauma Questionnaire.

### Group differences in CTQ scores

As shown in *Figure 1*, the PNES group had higher mean CTQ scores compared to epilepsy across all domains, in both the retrospective and prospective cohorts (see *Table 1*). In the retrospective cohort, these differences were large for the CTQ total score (*g* = 0.95, 95% CI = 0.46, 1.43), emotional abuse (*g* = 0.94, 95% CI = 0.47, 1.39), and emotional neglect (*g* = 0.87, 95% CI = 0.41, 1.32); medium sized differences were observed for physical abuse (*g* = 0.62, 95% CI = 0.14, 1.09), sexual abuse (*g* = 0.61, 95% CI = 0.14, 1.08), and physical neglect (*g* = 0.58, 95% CI = 0.13, 1.03). These findings were replicated in the prospective cohort for CTQ total score (*g* = 0.93, 95% CI = 0.28, 1.56), emotional abuse (*g* = 0.78, 95% CI = 0.16, 1.40), sexual abuse (*g* = 0.98, 95% CI = 0.32, 1.62), emotional neglect (*g* = 0.69, 95% CI = 0.09, 1.28), and physical neglect (*g* = 0.68, 95% CI = 0.06, 1.30), although the group difference for physical abuse was not replicated (*g* = 0.51, 95% CI = -0.09, 1.10).

**Figure 1.**
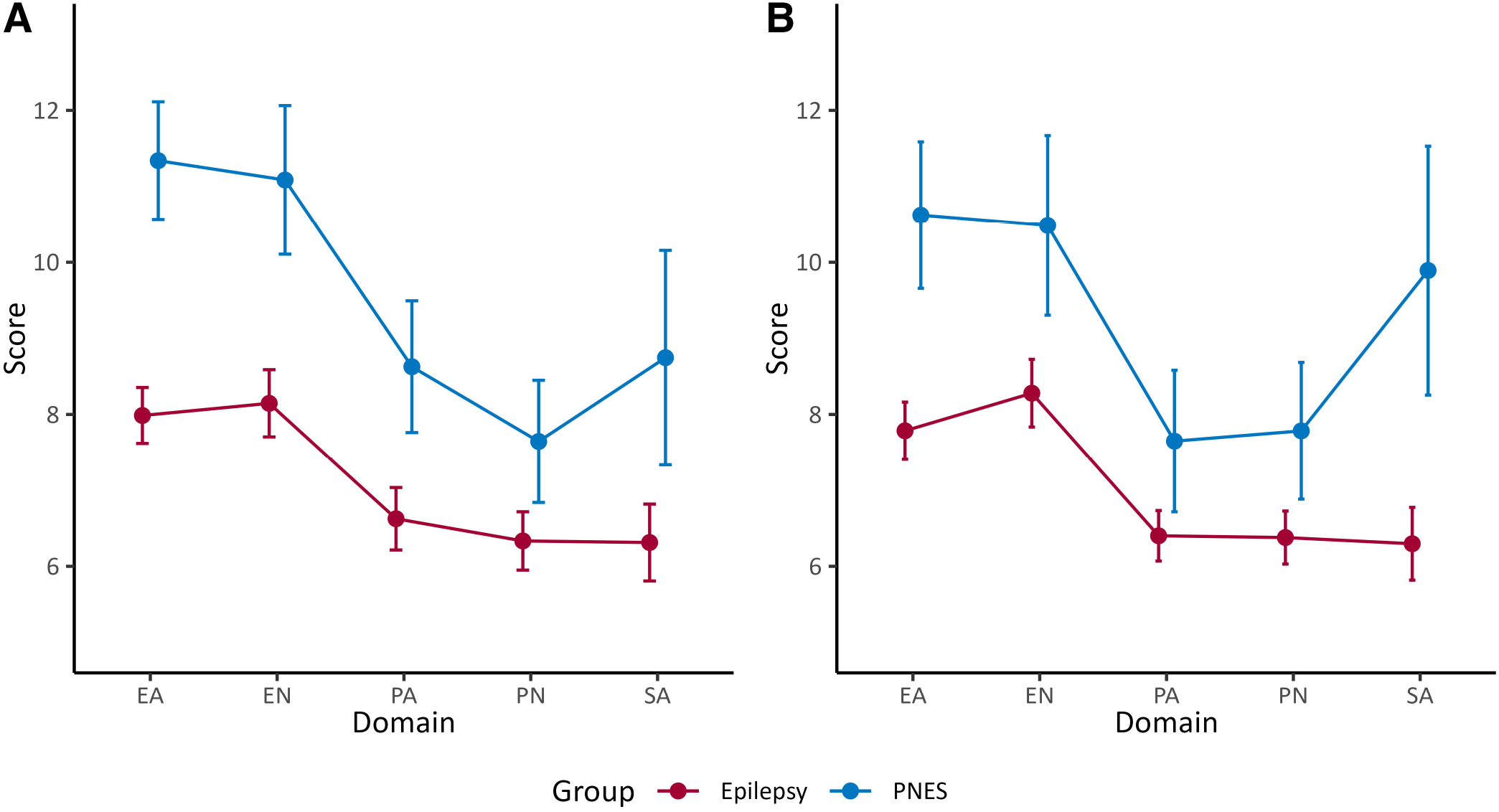
Means of CTQ domains scores across all domains in the retrospective (A) and prospective (B) cohorts. Error bars are 95% confidence intervals. Patients in the PNES group had higher mean scores across all domains, which was replicated in the prospective cohort except for physical abuse. In term of raw scores, the greatest differences appear to be in the EA, EN, and SA domains. Note: EA = emotional abuse, EN = emotional neglect, PA = physical abuse, PN = physical neglect, SA = sexual abuse.

### Analysis of trauma profiles

Generalised linear models (GLMMs) were computed to compare the CTQ profiles between the PNES and epilepsy groups. For the retrospective cohort there was a main effect for group (ω^2^_p_ = 0.09, 95% CI = 0.03, 0.17) but no evidence for an interaction between group and CTQ domain score (ω^2^_p_ = 0.00, 95% CI = -0.01, 0.01). The PNES group had higher scores than the epilepsy group when averaged across domains (β = 0.71, 95% CI = 0.41, 1.00), which is shown in *Figure 2*. This indicates that the CTQ profiles were uniformly elevated in PNES compared to epilepsy and that there was no evidence for elevations in specific CTQ domain scores. This profile is shown in *Figure 3*. These findings were replication in the prospective cohort, with evidence of a main effect o group (ω^2^_p_ = 0.07, 95% CI = 0.02, 0.14) but again no evidence for a group by domain (ω^2^_p_ = 0.00, 95% CI = -0.01, 0.01). The PNES group had higher scores than the epilepsy group when averaged across domains (β = 0.60, 95% CI = 0.25, 0.94).

**Figure 2.**
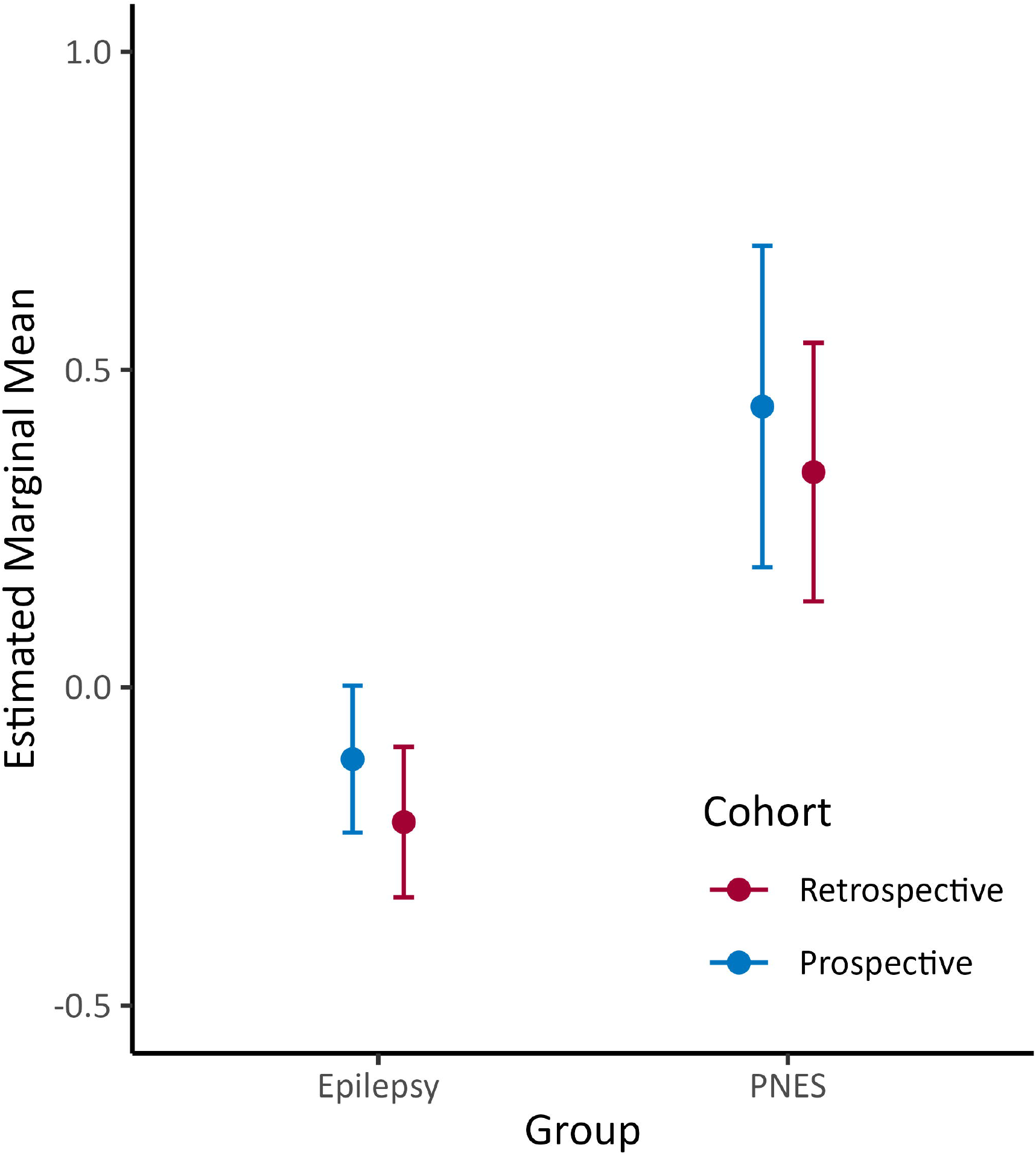
Estimated marginal means from the general linear mixed models (GLMM) showing overall higher CTQ scores (averaged across domains) in the PNES group compared to the epilepsy group. This finding was replicated in the prospective cohort.

**Figure 3.**
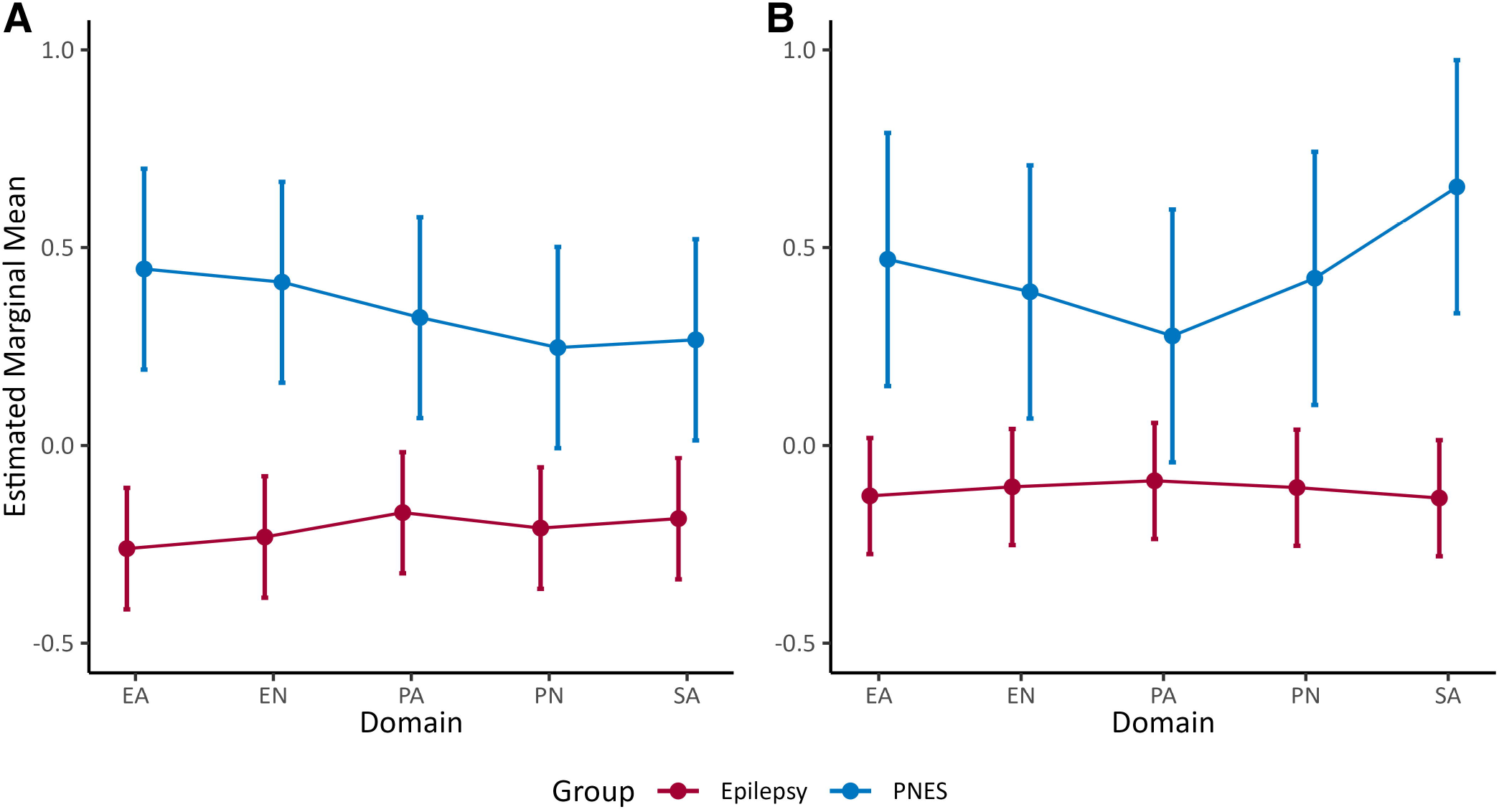
Estimated marginal means from the generalised linear models (GLMMs) comparing CTQ profiles across epilepsy and PNES groups. In the retrospective cohort (A) there was evidence for overall elevation in CTQ scores across domains in the PNES group compared to the epilepsy group. There was no evidence for a group by domain interaction, indicting a lack of evidence for elevations in specific domains. This finding was replicated in the prospective cohort (B). Note: EA = emotional abuse, EN = emotional neglect, PA = physical abuse, PN = physical neglect, SA = sexual abuse.

Additional analyses were computed to investigate the relationship between gender and CTQ domain scores. In both the retrospective and prospective cohorts there was no evidence for a main effect of gender, nor an interaction between gender and CTQ domain (all *p*s > .05). Statistically significant effects of gender did not emerge even when the PNES patients were considered in isolation after removal of the epilepsy patients from the analyses.

### *Classification of PNES and* epilepsy

Raw CTQ raw scores were analysed using receiver operating characteristic (ROC) curves for identification of PNES. As shown in *Table 2*, the CTQ total, emotional abuse, sexual abuse, emotional neglect, and physical neglect scores were able to classify cases as being PNES versus epilepsy to some degree, based on area under the curve (AUC) values that did not capture 0.50. Sensitivity was relatively low for these scores compared to specificity, resulting in higher negative predictive values (NPV) compared to positive predictive values (PPV).

**Table 2.**
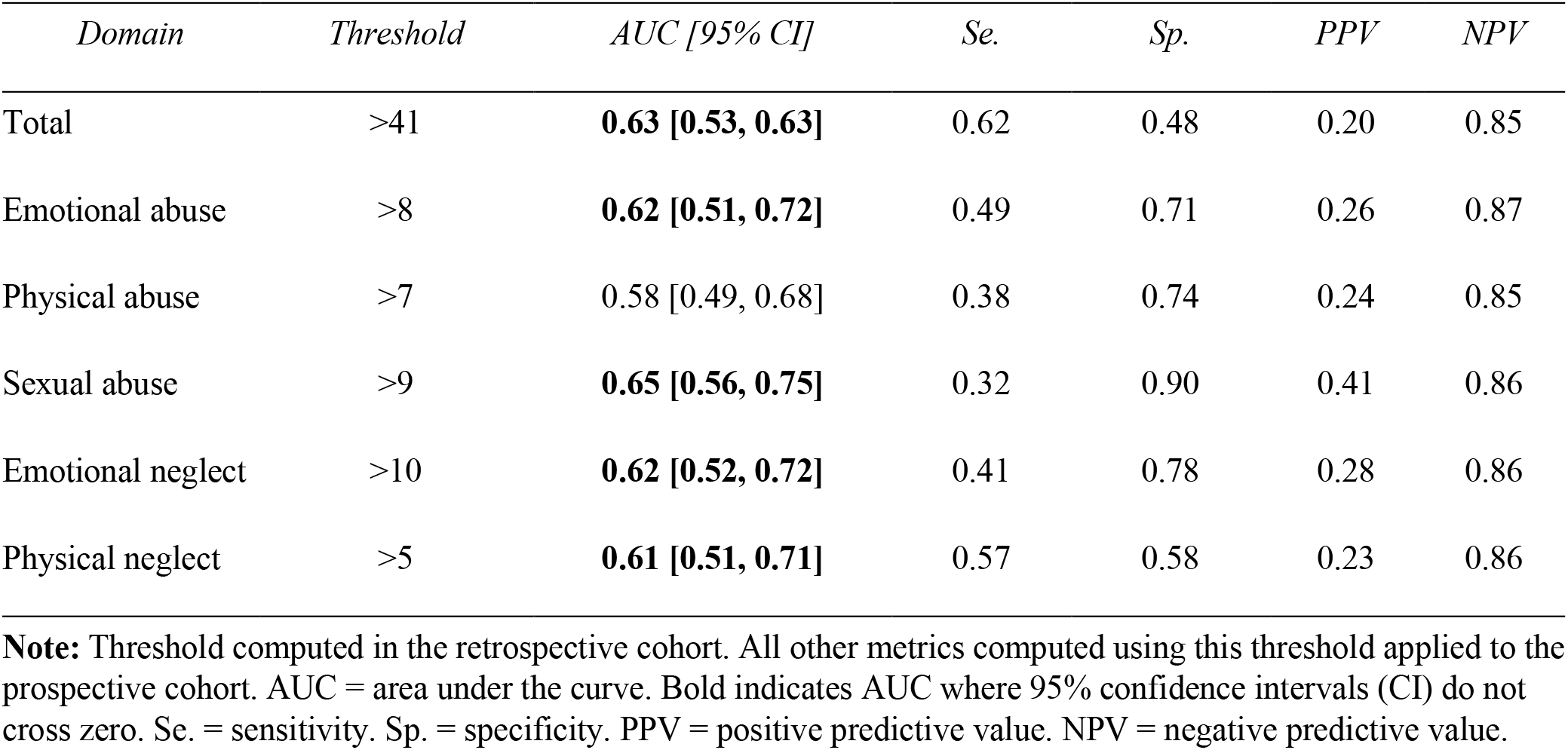
Raw classification performance of CTQ domain scores to identify PNES.

| *Domain* | *Threshold* | *AUC [95% CI]* | *Se.* | *Sp.* | *PPV* | *NPV* |
| --- | --- | --- | --- | --- | --- | --- |
| Total | >41 | **0.63 [0.53, 0.63]** | 0.62 | 0.48 | 0.20 | 0.85 |
| Emotional abuse | >8 | **0.62 [0.51, 0.72]** | 0.49 | 0.71 | 0.26 | 0.87 |
| Physical abuse | >7 | 0.58 [0.49, 0.68] | 0.38 | 0.74 | 0.24 | 0.85 |
| Sexual abuse | >9 | **0.65 [0.56, 0.75]** | 0.32 | 0.90 | 0.41 | 0.86 |
| Emotional neglect | >10 | **0.62 [0.52, 0.72]** | 0.41 | 0.78 | 0.28 | 0.86 |
| Physical neglect | >5 | **0.61 [0.51, 0.71]** | 0.57 | 0.58 | 0.23 | 0.86 |

**Note:** Threshold computed in the retrospective cohort. All other metrics computed using this threshold applied to the prospective cohort. AUC = area under the curve. Bold indicates AUC where 95% confidence intervals (CI) do not cross zero. Se. = sensitivity. Sp. = specificity. PPV = positive predictive value. NPV = negative predictive value.

Generalised linear models were then computed to investigate the relationship between CTQ scores and diagnostic group (PNES or epilepsy) above and beyond clinicodemographic data. This analysis was performed only in the prospective cohort, as clinicodemographic data were not reliably available in the retrospective cohort. As shown in *Table 3*, the baseline model was statistically significant (AIC = 145.49, Pseudo *R*^2^ = 0.36, *p* < .001). Only the number of antiseizure medications (ASM) was statistically supported, with the odds of having PNES compared to epilepsy reducing by 5 times for each prescribed ASM (OR = 0.20, 95% CI = [0.06, 0.43]). There was no evidence for an effect of age at admission (OR = 0.82, 95% CI = [0.50, 1.31]), disease duration (OR = 0.70, 95% CI = [0.31, 1.12]), gender (OR = 0.55, 95% CI = [0.16, 1.43]), seizure/event frequency (OR = 1.06, 95% CI = [0.70, 1.68]), or cognitive status (OR = 0.99, 95% CI = [0.59, 2.02]).

**Table 3.** Separate Generalised Linear Models (GZLMs) with CTQ scores.

| *Model* | *OR [95% CI]* | *AIC* | *Pseudo R<sup>2</sup>* | *Sig.* |
| --- | --- | --- | --- | --- |
| Baseline | - | 145.49 | 0.36 | < .001 |
| Total | **1.87 [1.26, 3.28]** | 139.70 | 0.41 | **.005** |
| Emotional abuse | **1.47 [1.03, 2.19]** | 144.14 | 0.38 | .07 |
| Physical abuse | 1.39 [0.88, 2.29] | 145.29 | 0.38 | .14 |
| Sexual abuse | **1.76 [1.16, 3.49]** | 140.23 | 0.41 | **.007** |
| Emotional neglect | 1.49 [1.03, 2.49] | 144.37 | 0.38 | .08 |
| Physical neglect | **2.02 [1.37, 3.70]** | 138.36 | 0.42 | **.003** |

**Note:** Results show the effect of adding each CTQ score to the baseline generalised linear model (GZLM). OR = odds ratio representing the increase in odds of having a PNES diagnosis for each standard deviation increase in the relevant CTQ score. AIC = Akaike information criteria. Sig. = statistical significance.

Several CTQ scores were associated with diagnostic group (PNES versus epilepsy) over and above clinicodemographic data. As shown in *Table 3*, marginal improvements in model fit were observed when the CTQ total, sexual abuse, and physical neglect scores were added to the model.

### Relationships between childhood trauma and clinicodemographic and psychopathology variables

As shown in *Table 4*, older age was weakly associated with higher scores on the physical neglect domain. Otherwise, there was no strong evidence for an association between childhood trauma, age, disease duration, seizure/event frequency, and number of prescribed ASMs. All CTQ domains were positively correlated with scores on the NDDI-E (depression), WDS (dissociation), PID-5 negative affect, PID-5 detachment, PID-5 disinhibition, and PID-5 psychoticism (r>.3).

**Table 4.** Associations Between Domain Scores, Clincodemographic, and Psychopathology scores.

| Variable | Emotional abuse | Emotional neglect | Sexual abuse | Physical abuse | Physical neglect |
| --- | --- | --- | --- | --- | --- |
| *Clinicodemographic* |  |  |  |  |  |
| Age | 0.05 | 0.10 | 0.08 | 0.10 | **0.25** |
| Disease duration | -0.03 | 0.07 | 0.08 | 0.07 | -0.01 |
| Event frequency | -0.11 | -0.03 | -0.12 | -0.04 | -0.03 |
| No. ASMs | -0.03 | -0.03 | -0.06 | -0.03 | 0.03 |
| *Psychometric* |  |  |  |  |  |
| GAD-7 | **0.36** | 0.14 | **0.19** | 0.14 | **0.27** |
| NDDI-E | **0.40** | **0.17** | **0.26** | **0.17** | **0.32** |
| NuCOG | 0.02 | 0.02 | 0.03 | 0.02 | -0.14 |
| WDS | **0.45** | **0.32** | **0.28** | **0.32** | **0.42** |
| PID-5 Negative affect | **0.42** | **0.31** | **0.17** | **0.31** | **0.24** |
| PID-5 Detachment | **0.33** | **0.24** | **0.19** | **0.24** | **0.37** |
| PID-5 Antagonism | 0.12 | **0.17** | 0.05 | **0.17** | 0.15 |
| PID-5 Disinhibition | **0.39** | **0.30** | **0.32** | **0.30** | **0.33** |
| PID-5 Psychoticism | **0.44** | **0.37** | **0.29** | **0.37** | **0.33** |

**Note:** Spearman's correlation coefficients are shown. Bold indicates coefficient for which the 95% confidence did not capture 0. GAD-7 = Generalized Anxiety Disorder-7; NDDI-E = Neurological Disorders Depression Inventory for Epilepsy; No. ASMs = GLM number of antiseizure medications; NuCOG = Neuropsychiatry Unit Cognitive Assessment; PID-5 = Personality Inventory for DSM-5; WDS = Wessex Dissociation Scale.

## Discussion

This study is the first to examine childhood trauma profiles between patients with PNES and epilepsy in two large samples of well-characterised patients undergoing investigation of seizure disorders. We found robust evidence that the overall level of reported childhood trauma was greater in patients diagnosed with PNES compared to epilepsy. PNES patients also reported greater emotional abuse, emotional neglect, physical neglect, and sexual abuse, but not physical abuse. These findings were consistent across both retrospective discovery and prospective validation cohorts. Overall childhood trauma, sexual abuse, and physical neglect were associated with diagnostic group (PNES versus epilepsy) over and above clinicodemographic factors. The severity of reported sexual abuse showed the best screening performance when used as an isolated measure, although it was not accurate enough for diagnostic or screening applications. The robustness of these findings is underscored by the similar results in the two cohorts examined in this study, both of which had a larger sample of patients with PNES than any previous literature in this area of research, which facilitated immediate replication of the findings. Additionally, all diagnoses were made by a multidisciplinary expert team after VEM that allowed a more rigorous stratification of our sample to include only patients with ‘pure’ PNES or epilepsy conditions.

Our study revealed new and robust findings that overall childhood trauma history is elevated in patients with PNES relative to those with epilepsy. Although previous studies have conducted similar research, those findings are inconsistent and less reliable due to methodological limitations of using imprecise diagnostic classifications (15) and smaller sample sizes (1, 41). Notably, the childhood trauma profile in our adults with PNES is remarkably consistent with the elevated childhood emotional abuse, emotional neglect, and sexual abuse found in an adolescent PNES patient group compared to healthy controls (42). The similar findings of multiple childhood trauma types closely associated with PNES aetiology, instead of earlier research only focusing on physical and sexual abuse, support the proposed mechanism of PNES whereby seizures manifest from abnormal psychological processes (6-9). PNES is highly comorbid with other psychopathologies, including depression, anxiety, and borderline personality disorders, and childhood trauma could be a common psychological underlying mechanism in both PNES and its common comorbid disorders (43-45). This interpretation is supported by the promising outcomes of PNES interventions targeting psychological wellbeing such as emotional processing and interpersonal relationship skills (46, 47). Recent evidence has also supported the efficacy of prolonged exposure therapy in PNES, which is a form of CBT that selectively addresses post-traumatic symptomatology (48). Future investigations of common psychological factors associated with PNES may further shed light on the aetiology of PNES.

Whether subjective severity of childhood trauma is pertinent to the risk of developing PNES has to date been unclear. Previous studies have only separately asserted that type or overall history of childhood trauma was predictive of later PNES diagnoses (11, 13, 14). Our findings that patients who experienced more severe childhood trauma are at higher risk of PNES regardless of the trauma subtypes, is supportive of the concept that rather than a specific childhood trauma type being predictive of later PNES diagnosis, subjective childhood trauma of any kind places patients at higher risk of PNES diagnosis than epilepsy in a linear fashion. It highlights the pertinence of trauma severity rather than the presence or absence of trauma when translating childhood trauma profiles into diagnostic strategies.

A limitation of the current study is the unbalanced gender ratio between PNES and epilepsy groups. It has been argued that the predominance of females diagnosed with PNES together with females being more susceptible to childhood emotional and sexual trauma might account for the higher reports of these trauma types in patients with PNES compared to patients with epilepsy (12). However, neither of the current cohorts showed any significant gender effect in childhood trauma profile, and logistic regression did not demonstrate any predictive effect of female gender on PNES diagnosis. Further exhaustive sensitivity analyses failed to find evidence for any gender effects or interaction. However, a large international study found that the gender ratio in the PNES population fluctuated across the lifespan, with female predominance in both adolescent-onset and adult-onset PNES populations but not in childhood-onset PNES (46). Moreover, the prevalence rate of childhood sexual abuse varies across different continents, with growing evidence suggesting higher rates in Australia than other regions such as Asia, Europe, and South America (47, 49). Hence, geographical differences in disclosures and/or occurrence of specific trauma types and overall childhood trauma may also limit the generalisability of our findings in other PNES populations. As such, future studies could explore whether the same childhood trauma profile revealed in the current study is consistent across patients with childhood-onset PNES and patients with different cultures, races, and ethnicities.

It is also important to note that both of our cohorts were recruited from inpatient VEM units. This is certainly a strength, as it minimises the probability of an incorrect clinical diagnosis. It does mean, however, that these findings do not necessarily generalise to the wider epilepsy or PNES populations. Further, the CTQ is a self-report measure of past trauma. As such, there are number of psychological factors at play (e.g., difficulty understanding questions, social desirability bias) that might result in a certain degree of bias. From the perspective of psychometric screening, such bias is less of a confounding factor. From the perspective of aetiological theory development, however, the effects of this bias are important to consider. Further research is certainly warranted in this regard.

We also tested the classification accuracy of using childhood trauma as a screening diagnostic test for PNES. While differences in trauma severity between PNES versus epilepsy showed solid statistical evidence of at least medium to large effect sizes from both cohorts, the sensitivity and specificity metrics for using childhood trauma as isolated measures for PNES screening were poor, which suggests that their use as psychometric screening instruments for PNES might be limited. At best, select measures showed high specificity and NPV, suggesting relatively high accuracy in ruling out a PNES diagnosis when patients report little childhood trauma experience. However, until these childhood trauma profiles are replicated in the broader literature, it is pragmatic to consider low childhood trauma profiles as a marker of low risk of PNES compared to epilepsy rather than having high diagnostic values per se.

To summarise, the pattern of elevated emotional abuse, emotional neglect, physical neglect, and sexual abuse found in patients with PNES supports the long-held concept that the experience of childhood trauma is a vulnerability factor for the development of PNES, with a history of more severe childhood trauma of any type raising the likelihood that seizures have a functional rather than epileptic basis. From a methodological point of view, current findings also support the further investigations of the CTQ as a routine screening tool for childhood trauma in these patient populations. Together these findings support the value of incorporating childhood trauma screening into routine clinical workflows and suggests that clinicians working in the epilepsy clinic should consider a differential diagnosis of PNES in patients with a severe childhood trauma history.

## Acknowledgments

No specific funding was received for this research.

## Author Contributions

**Tianren Yang:** Conceptualization, Data Curation, Formal Analysis, Investigation, Methodology, Project Administration, Writing – Original Draft Preparation. **Caitlin Roberts:** Conceptualization, Data Curation, Investigation, Methodology, Project Administration, Writing – Review & Editing. **Toby Winton-Brown:** Conceptualization, Investigation, Methodology, Project Administration, Writing – Review & Editing. **Michael Lloyd:** Conceptualization, Data Curation, Investigation, Methodology, Project Administration, Writing – Review & Editing. **Patrick Kwan:** Conceptualization, Investigation, Methodology, Project Administration, Writing – Review & Editing. **Terence J O’Brien:** Conceptualization, Investigation, Methodology, Project Administration, Writing – Review & Editing. **Dennis Velakoulis:** Conceptualization, Investigation, Methodology, Project Administration, Writing – Review & Editing. **Genevieve Rayner:** Conceptualization, Investigation, Methodology, Project Administration, Writing – Review & Editing. **Charles B Malpas:** Conceptualization, Investigation, Methodology, Project Administration, Formal Analysis, Writing – Review & Editing, Supervision.

